# Factors Associated with Adherence to Anti-Hypertensive Medication Among Patients Attending a Primary Health Centre in Ndola, Zambia: A Cross-Sectional Study

**DOI:** 10.64898/2026.09.11.26362862

**Authors:** Nancy Sikantongwe, Elliot Musonda, Florence Mabaso, Erick Sikantongwe, Elipilious Chibwaya, Chileleko Mapiki

## Abstract

**Background:** Hypertension is a leading modifiable risk factor for cardiovascular disease and premature death worldwide. Despite effective pharmacological therapy, poor adherence to anti-hypertensive medication remains the primary barrier to achieving blood pressure control, with non-adherence rates reaching 45-65% in sub-Saharan African settings. In Zambia, uncontrolled hypertension contributes substantially to hospital morbidity and mortality, yet context-specific data on adherence barriers remain limited. This study assessed factors associated with adherence to anti-hypertensive medication among patients attending Railway Surgery Urban Health Centre in Ndola, Zambia.

**Methods:** A cross-sectional study was conducted among 237 hypertensive patients attending Railway Surgery Urban Health Centre in Ndola. A structured, interviewer-administered questionnaire assessed socio-demographic characteristics, clinical factors, and medication adherence using the modified Hill-Bone Compliance Scale. Data were analysed using descriptive statistics, chi-square tests, and multivariable logistic regression (p<0.05).

**Results:** The overall adherence rate was 41.4%, with 58.6% of participants classified as non-adherent. The mean adherence score was 19.8 (SD=4.6) out of a possible 32. The most common reasons for non-adherence were running out of pills (mean=2.6), being away from home (mean=2.5), and forgetting to take medication (mean=2.4). Age (p=0.006), education level (p=0.001), and monthly income (p=0.001) were significantly associated with adherence. Clinical factors significantly associated with poorer adherence included presence of comorbidities (p=0.027), experience of side effects (p<0.001), and medication regimen complexity (p=0.007). Multivariable analysis identified age ≥51 years (AOR=2.89-3.24), secondary/tertiary education (AOR=2.84-3.56), monthly income ≥K1,000 (AOR=2.21-5.89), absence of comorbidities (AOR=0.51), absence of side effects (AOR=0.36), and simpler regimens (AOR=0.34) as factors independently associated with adherence. Health system barriers included long distances, reliance on public transport, missed appointments due to transport problems (35.4%), medication stock-outs (38.4%), and inadequate counselling time (49.8% received <5 minutes).

**Conclusion:** Adherence to anti-hypertensive medication at Railway Surgery Urban Health Centre is suboptimal, with multiple socio-demographic, clinical, and health system factors associated with non-adherence. Interventions should focus on regimen simplification, proactive side effect management, enhanced patient education, and addressing systemic barriers including medication supply and transport access.

## BACKGROUND

Hypertension poses a significant public health challenge globally, in Africa, and particularly in Zambia. The World Health Organization defines hypertension as having a systolic blood pressure of 140 mmHg or higher, or a diastolic blood pressure of 90 mmHg or above (World Health Organization, 2017). This condition often shows no symptoms, earning it the label of a “silent killer,” as people frequently remain unaware of their hypertension until serious health issues emerge (Yan et al., 2015; Rush et al., 2018).

Globally, hypertension affects nearly 1.28 billion individuals aged 30-79 years (World Health Organization, 2017). Cardiovascular diseases, to which hypertension significantly contributes, result in an estimated 17.9 million deaths annually, accounting for 32% of all deaths worldwide (Burnier & Egan, 2019; Abegaz et al., 2017). This situation is particularly concerning for low- and middle-income countries, as their already strained healthcare systems may become overwhelmed by increasing hypertension rates (De Geest et al., 2021).

Adherence to hypertension treatment is alarmingly low globally, with studies reporting that only about 50% of individuals with high blood pressure follow their prescribed antihypertensive regimens (Abegaz et al., 2017; Burnier & Egan, 2019). A systematic review and meta-analysis found that non-adherence to antihypertensive drugs remains a significant barrier to blood pressure control worldwide, with rates varying considerably across different settings and populations (Abegaz et al., 2017). In middle- to low-income countries, adherence can fall below 30% due to resource limitations, medication costs, and limited access to healthcare services (Kim et al., 2025; De Geest et al., 2021).

In Sub-Saharan Africa, the burden of hypertension is substantial, with approximately 27% of adults affected (Aminde et al., 2025; Chikumbu et al., 2024). Medication adherence among hypertensive patients in the region remains suboptimal, with estimates suggesting that only 30-50% of individuals consistently follow their prescribed treatment plans (Aminde et al., 2025; Egekeze et al., 2025). A meta-analysis of data from over 34,000 adults with hypertension in sub-Saharan Africa confirmed a high burden and increasing trend in non-adherence to blood pressure-lowering medications (Aminde et al., 2025). Health system barriers including medication stock-outs, long distances to health facilities, inadequate patient counseling, and high out-of-pocket costs contribute significantly to poor adherence in the region (Chikumbu et al., 2024; Egekeze et al., 2025).

In Zambia, hypertension prevalence is approximately 30% among adults (World Health Organization, 2017; Rush et al., 2018). The country witnesses an increase in hypertension-related complications, further straining its healthcare system (Muzambizi et al., 2021; Povia et al., 2023). Research on hypertension medication adherence in Zambia shows consistently suboptimal rates. A study at Matero Level One Hospital in Lusaka found that only 37.5% of hypertensive patients adhered to their medication regimen, with medication knowledge significantly affecting adherence (Kampamba et al., 2021). Mapiki et al. (2023), in their study of HIV patients on antihypertensive medication at Chanyanya Rural Health Center in Lusaka, reported an even lower adherence rate of 35.7%, highlighting the compounded challenges faced by patients with multiple chronic conditions. Mweene et al. (2010) identified multiple factors associated with poor adherence among hypertensive patients in Lusaka, including forgetfulness, side effects, and cost of medication. More recently, Povia et al. (2023) found that uncontrolled hypertension was associated with significant productivity losses and correlated with poor adherence and health system factors.

Several studies have identified key determinants of adherence to antihypertensive medication. A systematic literature review identified factors including age, education level, socioeconomic status, presence of comorbidities, side effects, and regimen complexity as significant predictors of adherence (Kim et al., 2025). In Zambia specifically, age, education, and income have been identified as factors influencing adherence (Kampamba et al., 2021; Mweene et al., 2010). The presence of comorbidities, particularly HIV, diabetes, and heart disease, has been associated with poorer adherence due to increased treatment burden and polypharmacy (Mapiki et al., 2023; Povia et al., 2023). Health system factors including medication availability, distance to health facilities, and patient-provider communication have also been identified as important determinants (Chikumbu et al., 2024; Muzambizi et al., 2021). Ngomas et al. (2022), in a study from neighboring Malawi, similarly found that medication side effects, cost, and health system barriers were significant contributors to non-adherence.

The Railway Surgery Urban Health Centre, an essential healthcare resource in Ndola, Zambia, has been grappling with a concerning trend. There has been an increase in both existing and new cases of hypertension at the clinic over recent months, highlighting a significant public health concern. Preliminary data from the clinic’s Outpatient Department Hypertension Book (2025) reveal an alarming pattern: between November 2024 and January 2025, there has been a marked increase in both new hypertension diagnoses and existing patients presenting with persistently uncontrolled blood pressure. In November 2024, there were 50 reported hypertension cases, with 30 being known cases and 20 new ones. In December 2024, that number included 42 known cases and 30 fresh diagnoses, totaling 72 cases. By January 2025, this comprised 36 new cases and 55 established cases, raising the cumulative total to 91. This troubling rise in hypertension cases underscores the urgent need for research into factors that influence patient adherence to antihypertensive treatments.

Given these findings, it is important to investigate the underlying reasons for the low adherence to antihypertensive medication among patients at the Railway Surgery Urban Health Centre. This study, therefore, sought to assess the factors influencing adherence to anti-hypertensive medication among patients attending Railway Surgery Urban Health Centre in Ndola, Zambia. The findings will provide the evidence base required to develop targeted, feasible, and context-appropriate interventions to improve adherence, enhance clinical outcomes, and curb the rising tide of uncontrolled hypertension at this facility.

## METHODS

### Study Design and Setting

This cross-sectional study was conducted with data collected from July to September 2025 at Railway Surgery Urban Health Centre in Ndola, Zambia. Railway Surgery Urban Health Centre is a public (government-run) primary healthcare facility located near downtown Ndola. The centre provides outpatient services, including hypertension management, to local residents. The facility was selected due to the preliminary observation of an increase in hypertension cases and concerns about suboptimal blood pressure control among patients on antihypertensive therapy.

### Study Population

The study population comprised patients diagnosed with hypertension and attending Railway Surgery Urban Health Centre for routine care. The target population included patients who were 18 years and above, diagnosed with hypertension, and currently prescribed antihypertensive medication.

### Inclusion and Exclusion Criteria

Patients were eligible for inclusion if they had a confirmed diagnosis of hypertension, were currently prescribed antihypertensive medication, were aged 18 years or above, attended Railway Surgery Urban Health Centre in Ndola, Zambia, and were willing and able to provide informed consent. Patients were excluded from the study if they had no confirmed diagnosis of hypertension, were under 18 years of age, had severe cognitive impairments or mental deficiency preventing effective communication, were unable to communicate effectively with the investigator, or were participating in another clinical trial or study. Patients with comorbidities including diabetes, HIV/AIDS, and heart disease were not excluded, as these conditions were examined as independent variables in the analysis.

### Sample Size Determination

The sample size was calculated using Cochran’s formula for cross-sectional studies: n = [Z² × p × (1-p)] / e², where Z = 1.96 (95% confidence level), p = 0.19 (estimated proportion based on hypertension prevalence from the Zambia WHO STEPS Survey, 2017), and e = 0.05 (5% margin of error). The calculation yielded: n = [(1.96)² × 0.19 × (0.81)] ÷ (0.05)² = [3.8416 × 0.1539] ÷ 0.0025 = 0.5912 ÷ 0.0025 = 236.48, which was rounded to 237 participants. The use of p=0.19 from the WHO STEPS Survey (2017) was justified as it represented the best available estimate of hypertension prevalence in the Zambian adult population at the time of study design, providing a conservative sample size estimate that would be adequate for detecting associations between multiple independent variables and adherence status.

### Sampling and Recruitment Procedure

Simple random sampling was employed to select participants from the clinic’s patient register. Each eligible patient was assigned a unique number, and a random number generator was used to select the required number of participants, ensuring that every qualified patient had an equal opportunity of being selected. A research assistant approached selected patients in a private area of the clinic, provided detailed information about the study including its purpose, procedures, potential risks and benefits, and obtained written informed consent from those who agreed to participate. Patients who declined to participate were recorded but not replaced, and the final sample consisted of all consenting participants.

### Data Collection Tools

A structured questionnaire was developed as the primary data collection tool, consisting of four sections aligned with the study objectives. Section A captured socio-demographic factors including age, sex, education level, employment status, marital status, and monthly income. Section B gathered clinical information including duration since hypertension diagnosis, presence of comorbidities, type of comorbidity, experience of medication side effects, type of side effects, and number of pills taken per day. Section C employed the modified Hill-Bone Compliance Scale to measure adherence levels, adapted from Mweene et al. (2010). The modified Hill-Bone Compliance Scale consisted of nine items covering three sub-dimensions: medication taking behaviour (four items), appointment keeping (three items), and medication refill practices (two items). Each item was rated on a four-point Likert scale ranging from 1 (none of the time) to 4 (all of the time). Total scores ranged from 8 to 32, with higher scores indicating poorer adherence. For analysis, adherence was categorized as adherent (score ≤16) or non-adherent (score >16), consistent with the scoring approach used by Mweene et al. (2010). Section D collected information on health system factors including distance from clinic, mode of transport, missed appointments due to transport problems, medication stock-outs in the past six months, counselling time, and provider explanation of medication.

### Validity and Reliability

The questionnaire was developed based on an extensive literature review and the conceptual framework adapted from the World Health Organization’s Multidimensional Model of Adherence. The tool was reviewed by research supervisors and experienced clinicians to ensure content validity and comprehensiveness. The Hill-Bone scale has established construct validity through numerous studies demonstrating its ability to distinguish between groups with known differences in adherence behaviour (Mweene et al., 2010). The modified Hill-Bone Compliance Scale demonstrated acceptable internal consistency in this study, with a Cronbach’s alpha coefficient of 0.78, indicating good reliability.

The questionnaire was pre-tested on 15 hypertensive patients at Railway Surgery Urban Health Centre to assess clarity, flow, and comprehension. Ambiguous questions were revised based on pre-test findings. The questionnaire was developed in English and then translated into the local language (Bemba) using a forward-backward translation method. Two independent bilingual translators performed the forward translation, and a third translator reconciled any discrepancies. A different set of translators performed the backward translation to verify semantic equivalence with the original English version.

### Data Collection Methods

Face-to-face interviewer-administered questionnaires were used to collect data. Prior to data collection, ethical approval was obtained from the Chreso University Research Ethics Committee (CUREC) and permission was secured from the clinic management at Railway Surgery Urban Health Centre. On each data collection day, potential participants were identified from the clinic’s patient register. Simple random sampling was used to select eligible patients until the sample size was achieved. Eligible patients were approached in a private area, provided with detailed information about the study, and given an opportunity to ask questions. Those who agreed to participate signed or thumb-printed a consent form.

Research assistants were trained for two days on the study objectives, questionnaire administration, ethical considerations, and maintaining confidentiality. Training included role-play sessions and observation of initial interviews to ensure consistency in question delivery and response recording. Completed questionnaires were reviewed daily for completeness and consistency, with any errors or omissions addressed through immediate follow-up where possible.

### Variables and Definitions

The primary outcome variable was adherence to anti-hypertensive medication, measured using the modified Hill-Bone Compliance Scale and dichotomized as adherent (score ≤16) or non-adherent (score >16). Independent variables included socio-demographic factors (age categorized as 18-35, 36-50, 51-65, above 65 years; sex; marital status; education level; employment status; monthly income), clinical factors (duration of hypertension diagnosis; presence of comorbidities; type of comorbidity; experience of side effects; type of side effect; number of pills per day), and health system factors (distance from clinic; mode of transport; missed appointments due to transport; medication stock-outs in past 6 months; counselling time; provider explanation; monthly expenditure on medication and transport).

### Statistical Analysis

Data were entered into Microsoft Excel and analysed using IBM SPSS Statistics version 26. Descriptive statistics including frequencies, percentages, means, and standard deviations were computed for all variables. For bivariate analysis, chi-square tests examined associations between categorical variables and adherence categories, with Fisher’s exact test used where expected cell counts were below five. The strength of associations was assessed using odds ratios with 95% confidence intervals. Variables with p<0.10 in bivariate analysis were considered for inclusion in the multivariable model.

For multivariable analysis, a backward stepwise logistic regression approach was employed. This method begins with all candidate variables entered into the model simultaneously and then sequentially removes those that do not contribute significantly to the model’s predictive ability, based on the likelihood ratio test. This approach was chosen because it balances the need to control for potential confounders while maintaining model parsimony, which is particularly important given the sample size and the need to avoid overfitting. The number of variables initially entered was limited to ensure an adequate events-per-variable (EPV) ratio of at least 10 events per variable, with the final model retaining only variables with p<0.05. Adjusted odds ratios with 95% confidence intervals were reported. Model fit was assessed using the Hosmer-Lemeshow goodness-of-fit test. Multicollinearity was assessed using variance inflation factors, with values less than 5 considered acceptable. Missing data were handled by complete case analysis, as the proportion of missing data was less than 3%.

### Ethical Considerations

Ethical approval was obtained from the Chreso University Research Ethics Committee (CUREC) [Approval ref number: 22321-05-2025.]. Permission was also obtained from the clinic management at Railway Surgery Urban Health Centre. All participants provided written informed consent. For participants who could not read or write, consent was obtained through a witness who confirmed that the information was explained in a language the participant understood, and the participant provided a thumbprint. Participants were fully informed of the study’s purpose, procedures, potential risks and benefits, and their right to withdraw at any time without consequences. Privacy and confidentiality were maintained during questionnaire administration; interviews were conducted in private settings within the clinic. Data were anonymized using unique participant codes and stored securely with password protection, with access restricted to the research team. Participants who expressed health concerns, reported side effects, or requested additional information about their hypertension management were referred to the clinic’s healthcare providers for appropriate care.

## RESULTS

### Participant Flow and Response Rate

A total of 255 eligible patients were identified from the clinic’s patient register and invited to participate in the study. Of these, 237 patients consented and completed the questionnaire, yielding a response rate of 93.0%. Eighteen patients declined to participate, citing reasons including lack of time (n=10), disinterest in the study (n=5), and feeling unwell on the day of recruitment (n=3). No participants were excluded after consenting, and all 237 completed questionnaires were included in the final analysis. There were no missing data for the primary outcome variables; missing data for any variable were minimal (<3%) and were handled by complete case analysis.

### Demographic Characteristics

Table 1 summarizes the demographic characteristics of participants. The majority of participants (36.3%) were aged between 51-65 years, with a mean age of 54.2 years (SD=12.8). Females comprised a larger proportion of the study population (57.0%), and most participants were married (62.4%). Regarding education, the largest proportion had primary school education (32.9%). Self-employed individuals formed the largest employment group (28.3%), followed by the unemployed (24.5%). Concerning monthly income, the largest proportion (30.8%) reported having no income.

**Table 1:** Distribution of Participants by Socio-Demographic Characteristics.

| Variable | Category | Frequency (n) | Percentage (%) |
| --- | --- | --- | --- |
| **Age (years)** | 18-35 | 28 | 11.8 |
|  | 36-50 | 79 | 33.3 |
|  | 51-65 | 86 | 36.3 |
|  | Above 65 | 44 | 18.6 |
|  | **Total** | **237** | **100** |
| **Sex** | Male | 102 | 43.0 |
|  | Female | 135 | 57.0 |
|  | **Total** | **237** | **100** |
| **Marital Status** | Married | 148 | 62.4 |
|  | Single | 31 | 13.1 |
|  | Divorced/Separated | 24 | 10.1 |
|  | Widowed | 34 | 14.4 |
|  | **Total** | **237** | **100** |
| **Education Level** | No formal education | 43 | 18.1 |
|  | Primary school | 78 | 32.9 |
|  | Secondary school | 69 | 29.1 |
|  | Tertiary/College/University | 47 | 19.9 |
|  | **Total** | **237** | **100** |
| **Employment Status** | Employed (formal sector) | 52 | 21.9 |
|  | Self-employed | 67 | 28.3 |
|  | Unemployed | 58 | 24.5 |
|  | Retired | 45 | 19.0 |
|  | Student | 15 | 6.3 |
|  | **Total** | **237** | **100** |
| **Monthly Income** | No income | 73 | 30.8 |
|  | Less than K1,000 | 62 | 26.2 |
|  | K1,000 - K2,999 | 51 | 21.5 |
|  | K3,000 - K4,999 | 31 | 13.1 |
|  | K5,000 and above | 20 | 8.4 |
|  | **Total** | **237** | **100** |

### Clinical Characteristics

Table 2 presents the clinical characteristics of participants. The majority of participants (37.6%) had been diagnosed with hypertension for 1-5 years. Comorbidities were present in 45.6% of participants. Among those with comorbidities, diabetes was the most common (53.7%), followed by heart disease (29.6%). Regarding medication side effects, 40.9% of participants reported experiencing side effects from their anti-hypertensive medication. Nearly half of participants (47.3%) took two pills per day, while 28.2% took three or more pills daily.

**Table 2:** Distribution of Participants by Clinical Characteristics.

| Characteristic | Category | Frequency (n) | Percentage (%) |
| --- | --- | --- | --- |
| **Duration of Hypertension** | Less than 1 year | 31 | 13.1 |
|  | 1 - 5 years | 89 | 37.6 |
|  | 6 - 10 years | 72 | 30.4 |
|  | More than 10 years | 45 | 18.9 |
|  | **Total** | **237** | **100** |
| **Presence of Comorbidities** | Yes | 108 | 45.6 |
|  | No | 129 | 54.4 |
|  | **Total** | **237** | **100** |
| **Type of Comorbidity (n=108)** | Diabetes | 58 | 53.7 |
|  | Heart disease | 32 | 29.6 |
|  | Kidney disease | 11 | 10.2 |
|  | Other | 7 | 6.5 |
|  | **Total** | **108** | **100** |
| **Experience of Side Effects** | Yes | 97 | 40.9 |
|  | No | 140 | 59.1 |
|  | **Total** | **237** | **100** |
| **Number of Pills Per Day** | 1 pill | 58 | 24.5 |
|  | 2 pills | 112 | 47.3 |
|  | 3 or more pills | 67 | 28.2 |
|  | **Total** | **237** | **100** |

**Table 3:** Distribution of Adherence Scores.

| Adherence Category | Frequency (n) | Percentage (%) |
| --- | --- | --- |
| Adherent (Score ≤ 16) | 98 | 41.4 |
| Non-adherent (Score > 16) | 139 | 58.6 |
| **Total** | **237** | **100** |

### Level of Adherence to Anti-Hypertensive Medication

Adherence was measured using the modified Hill-Bone Compliance Scale, with scores ranging from 8 to 32. Lower scores indicate better adherence. The mean adherence score was 19.8 (SD=4.6).

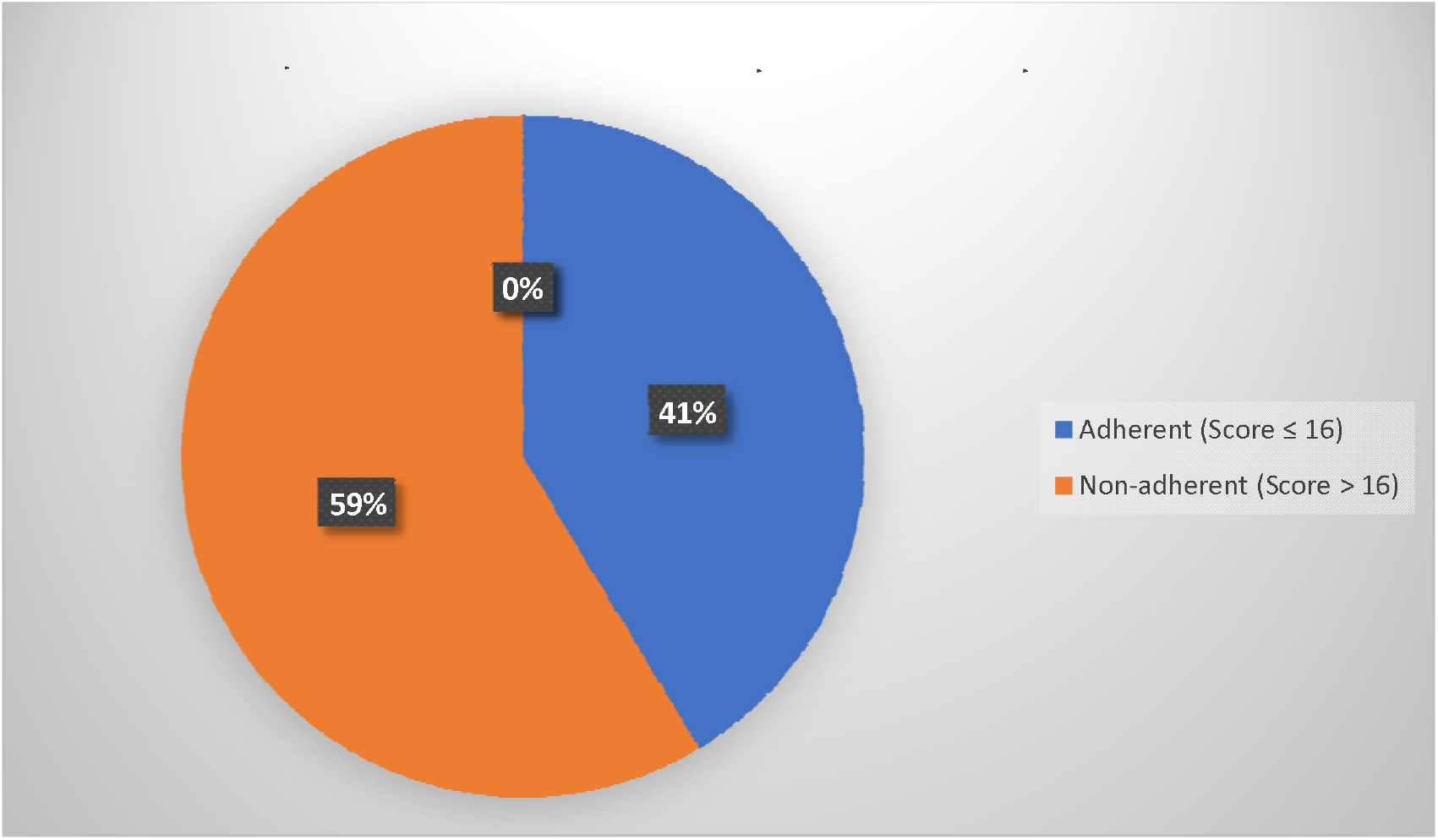

The overall adherence rate was 41.4%, with 58.6% of participants classified as non-adherent. Analysis of individual items revealed that the most common reasons for non-adherence were running out of pills (mean=2.6), being away from home (mean=2.5), and forgetting to take medication (mean=2.4). The least common reason was deciding not to take medication (mean=1.8), suggesting that most non-adherence was unintentional rather than intentional.

### Association Between Socio-Demographic Factors and Adherence

Chi-square analysis examined associations between socio-demographic variables and adherence categories (Table 4). Three socio-demographic factors showed statistically significant associations with medication adherence: age (p=0.006), education level (p=0.001), and monthly income (p=0.001). Adherence rates increased progressively with age, from 21.4% among those aged 18-35 years to 50.0% among those aged above 65 years. Adherence rates also increased with higher education levels, from 23.3% among those with no formal education to 57.4% among those with tertiary education. Similarly, adherence rates increased with income level, from 27.4% among those with no income to 75.0% among those earning K5,000 and above. Sex (p=0.632), marital status (p=0.409), and employment status (p=0.177) did not show statistically significant associations with adherence.

**Table 4:**
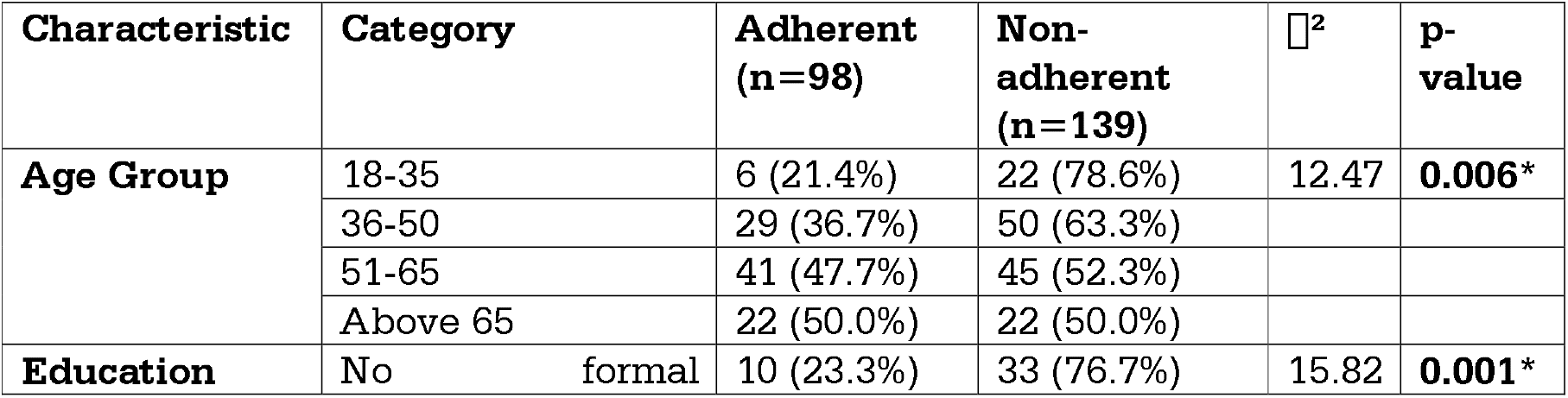
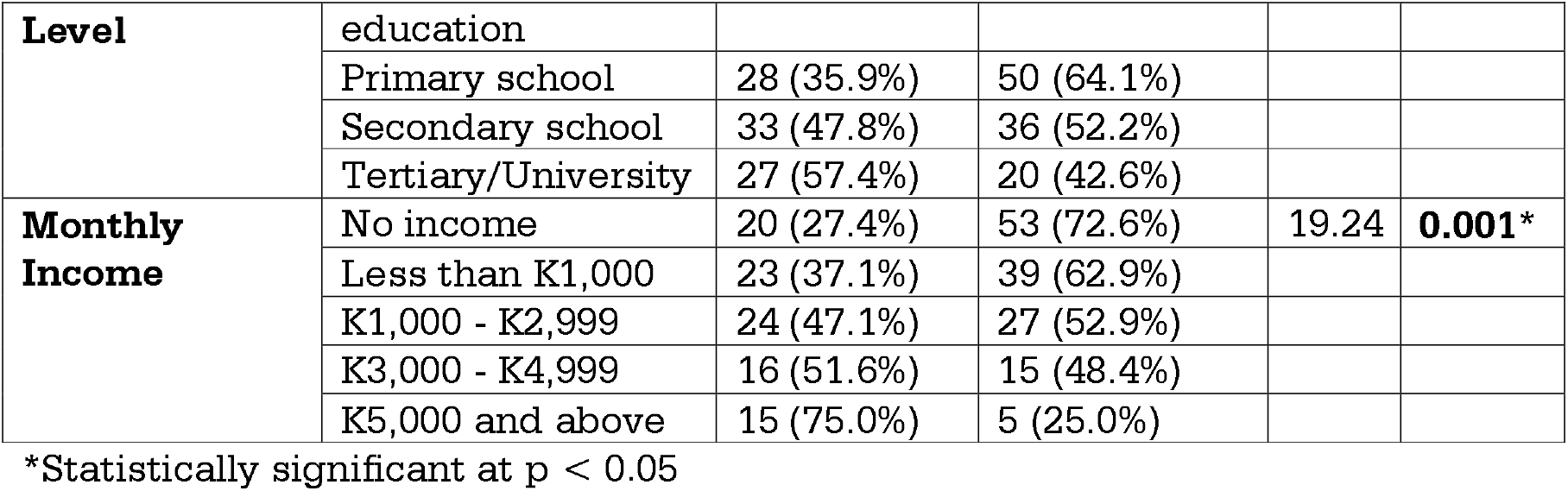
Association Between Socio-Demographic Factors and Adherence.

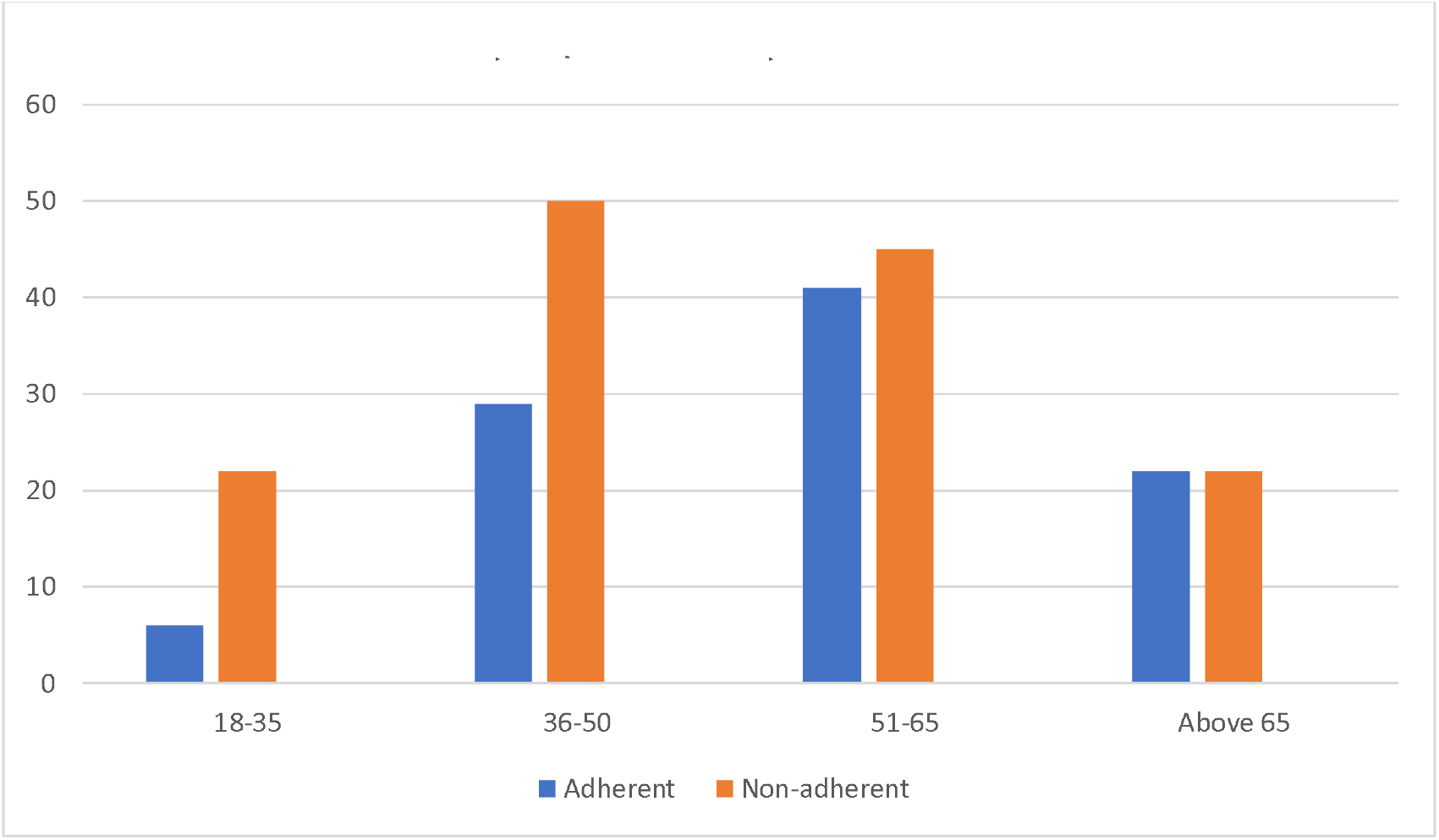

### Association Between Clinical Factors and Adherence

Chi-square tests examined associations between clinical factors and adherence categories (Table 5). Side effects showed a strong association with adherence (p<0.001); patients who experienced side effects had significantly lower adherence rates (26.8%) compared to those who did not (51.4%). There was a significant association between having comorbidities and adherence (p=0.027); patients with comorbidities had lower adherence rates (33.3%) compared to those without (48.1%). Among specific comorbidities, diabetes (p=0.042) and heart disease (p=0.025) were significantly associated with poorer adherence. The number of pills taken per day was significantly associated with adherence (p=0.007); adherence rates decreased from 55.2% for those taking one pill to 29.9% for those taking three or more pills daily. Duration of hypertension diagnosis (p=0.669) did not show a statistically significant association with adherence.

**Table 5:** Association Between Clinical Factors and Adherence.

| Characteristic | Category | Adherent (n=98) | Non-adherent (n=139) | $\chi^2$ | p-value |
| --- | --- | --- | --- | --- | --- |
| **Presence of Comorbidities** | Yes | 36 (33.3%) | 72 (66.7%) | 4.89 | **0.027\*** |
|  | No | 62 (48.1%) | 67 (51.9%) |  |  |
| **Experience of Side Effects** | Yes | 26 (26.8%) | 71 (73.2%) | 14.67 | **<0.001\*** |
|  | No | 72 (51.4%) | 68 (48.6%) |  |  |
| **Number of Pills Per Day** | 1 pill | 32 (55.2%) | 26 (44.8%) | 9.83 | **0.007\*** |
|  | 2 pills | 46 (41.1%) | 66 (58.9%) |  |  |
|  | 3 or more pills | 20 (29.9%) | 47 (70.1%) |  |  |

\*Statistically significant at $p < 0.05$

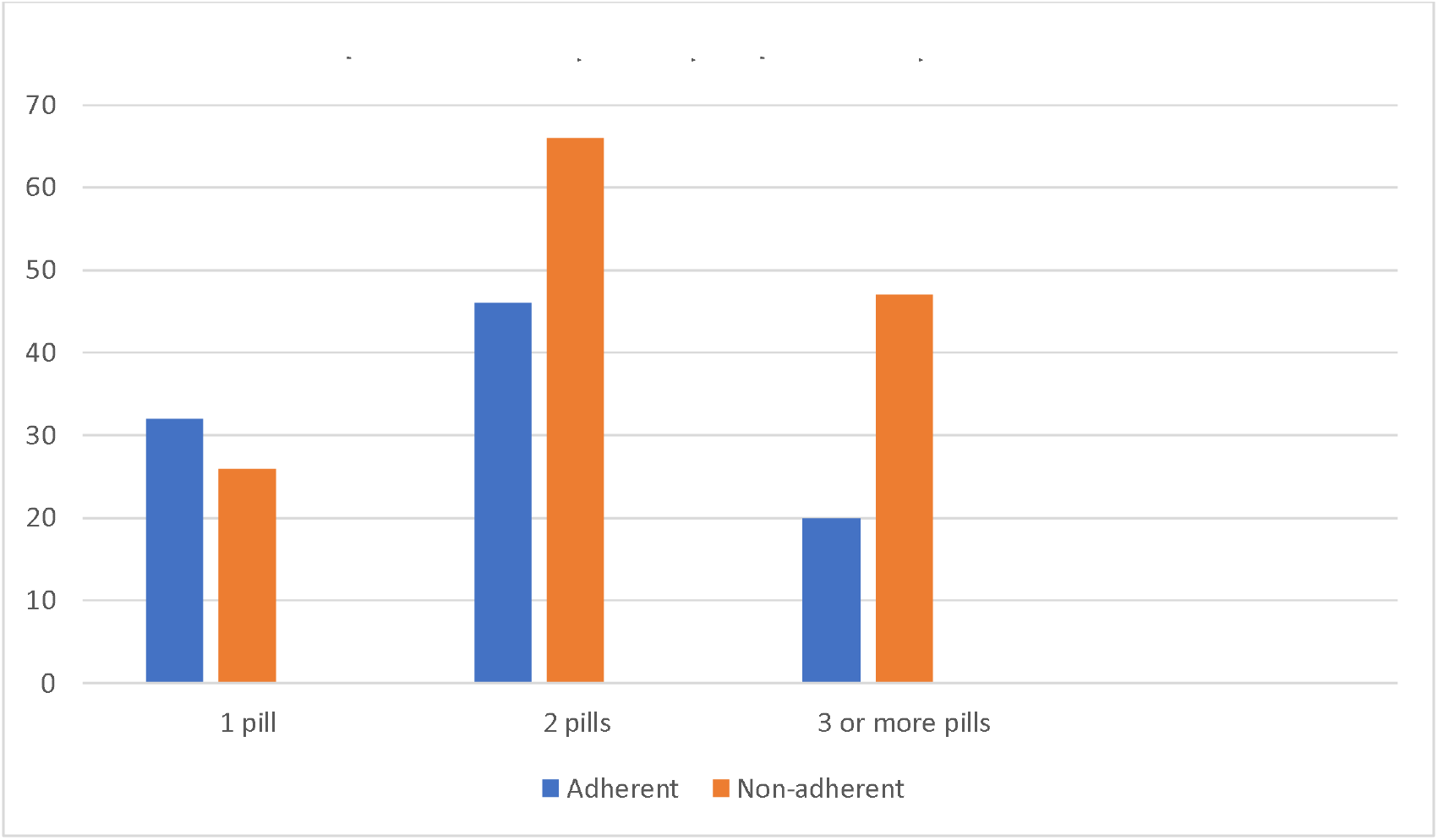

### Multivariable Analysis: Independent Predictors of Adherence

Variables with p<0.10 in bivariate analysis (age, education level, monthly income, presence of comorbidities, experience of side effects, and number of pills per day) were entered into a multivariable logistic regression model. A backward stepwise elimination approach was employed, beginning with all six variables in the model. Variables were removed sequentially based on the likelihood ratio test, with removal criteria set at p>0.05. The final model retained all six variables, as each contributed significantly to the model’s predictive ability. The events-per-variable (EPV) ratio was 16.3 (98 adherent patients divided by 6 variables), exceeding the recommended minimum of 10 events per variable.

The final model demonstrated good fit, with a Hosmer-Lemeshow test statistic of □²=8.42 (df=8, p=0.393), indicating no significant deviation between observed and predicted values. The model explained 28% of the variance in adherence (Nagelkerke R²=0.28) and demonstrated acceptable discrimination with a c-statistic (AUC) of 0.76. Multicollinearity was assessed using variance inflation factors, with all values below 2.0, indicating no significant multicollinearity.

The logistic regression analysis identified six independent predictors of adherence. Patients aged 51-65 years were nearly three times more likely to be adherent (AOR=2.89, 95% CI: 1.12-7.46), and those above 65 years were more than three times as likely to be adherent (AOR=3.24, 95% CI: 1.18-8.91) compared to those aged 18-35 years. Patients with secondary education were nearly three times more likely to be adherent (AOR=2.84, 95% CI: 1.28-6.31), while those with tertiary education were more than three and a half times more likely to be adherent (AOR=3.56, 95% CI: 1.45-8.74) compared to those with no formal education. Higher income was strongly associated with adherence; patients earning K1,000-K2,999 were twice as likely to be adherent (AOR=2.21, 95% CI: 1.03-4.74), while those earning K5,000 and above were nearly six times more likely to be adherent (AOR=5.89, 95% CI: 1.92-18.07) compared to those with no income. Patients with comorbidities were 49% less likely to be adherent compared to those without (AOR=0.51, 95% CI: 0.29-0.89). Patients who experienced side effects were 64% less likely to be adherent compared to those who did not (AOR=0.36, 95% CI: 0.20-0.65). Patients taking three or more pills daily were 66% less likely to be adherent compared to those taking one pill daily (AOR=0.34, 95% CI: 0.17-0.68).

### Health System Factors

Although not a specific objective, health system factors were assessed to provide context for adherence behaviour (Table 7). The majority of participants (41.3%) lived 5-10 km from the clinic, while 21.1% lived more than 10 km away. Most participants (59.9%) relied on public transport to reach the clinic. Over one-third (35.4%) reported having missed a clinic appointment due to lack of transport. Medication stock-outs were reported by 38.4% of participants in the past six months. Regarding patient-provider interaction, nearly half (49.8%) reported that counselling time was less than five minutes, and 19.4% reported receiving no counselling at all. Only 34.6% felt that healthcare providers always explained things in a way they could understand. The mean monthly expenditure on medication and transport was K187.50 (SD=94.30).

**Table 6:**
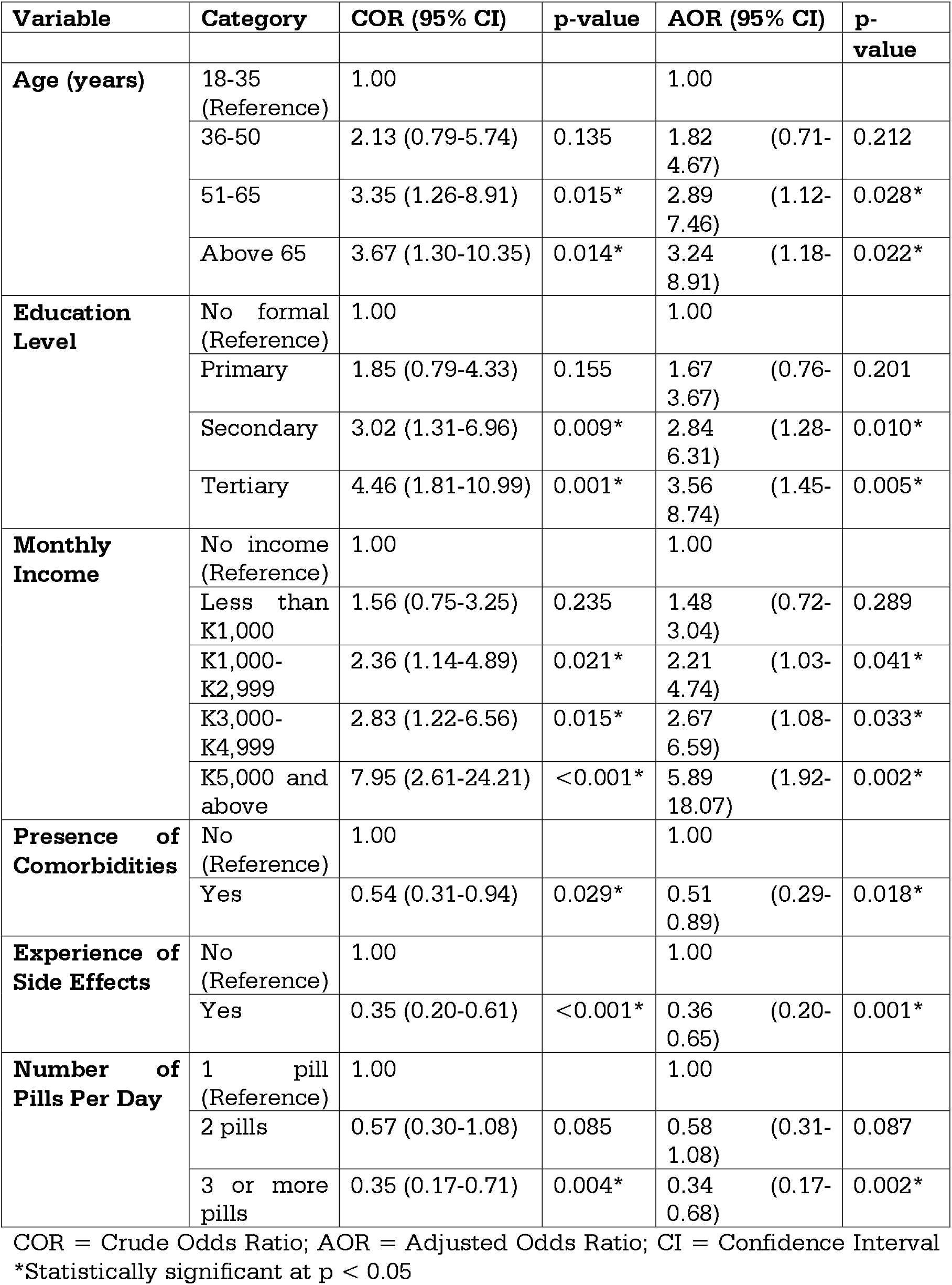
Bivariate and Multivariable Logistic Regression Analysis of Factors Associated with Adherence.

**Table 7:** Health System Factors.

| Factor | Category | Frequency (n) | Percentage (%) |
| --- | --- | --- | --- |
| Distance from Clinic | Less than 5 km | 89 | 37.6 |
|  | 5 - 10 km | 98 | 41.3 |
|  | More than 10 km | 50 | 21.1 |
|  | **Total** | **237** | **100** |
| Mode of Transport | Walking | 67 | 28.3 |
|  | By bus/taxi | 142 | 59.9 |
|  | By private car | 28 | 11.8 |
|  | **Total** | **237** | **100** |
| Missed Appointment Due to Transport | Yes | 84 | 35.4 |
|  | No | 153 | 64.6 |
|  | **Total** | **237** | **100** |
| Medication Stock-out in Past 6 Months | Yes | 91 | 38.4 |
|  | No | 146 | 61.6 |
|  | **Total** | **237** | **100** |
| Counselling Time | Less than 5 minutes | 118 | 49.8 |
|  | 5 minutes or more | 73 | 30.8 |
|  | Did not receive counselling | 46 | 19.4 |
|  | **Total** | **237** | **100** |
| Explanation by Providers | Always | 82 | 34.6 |
|  | Sometimes | 89 | 37.6 |
|  | Rarely | 41 | 17.3 |
|  | Never | 25 | 10.5 |
|  | **Total** | **237** | **100** |
| **Mean Monthly Expenditure on Medication and Transport** |  | **K187.50** | **(SD=94.30)** |

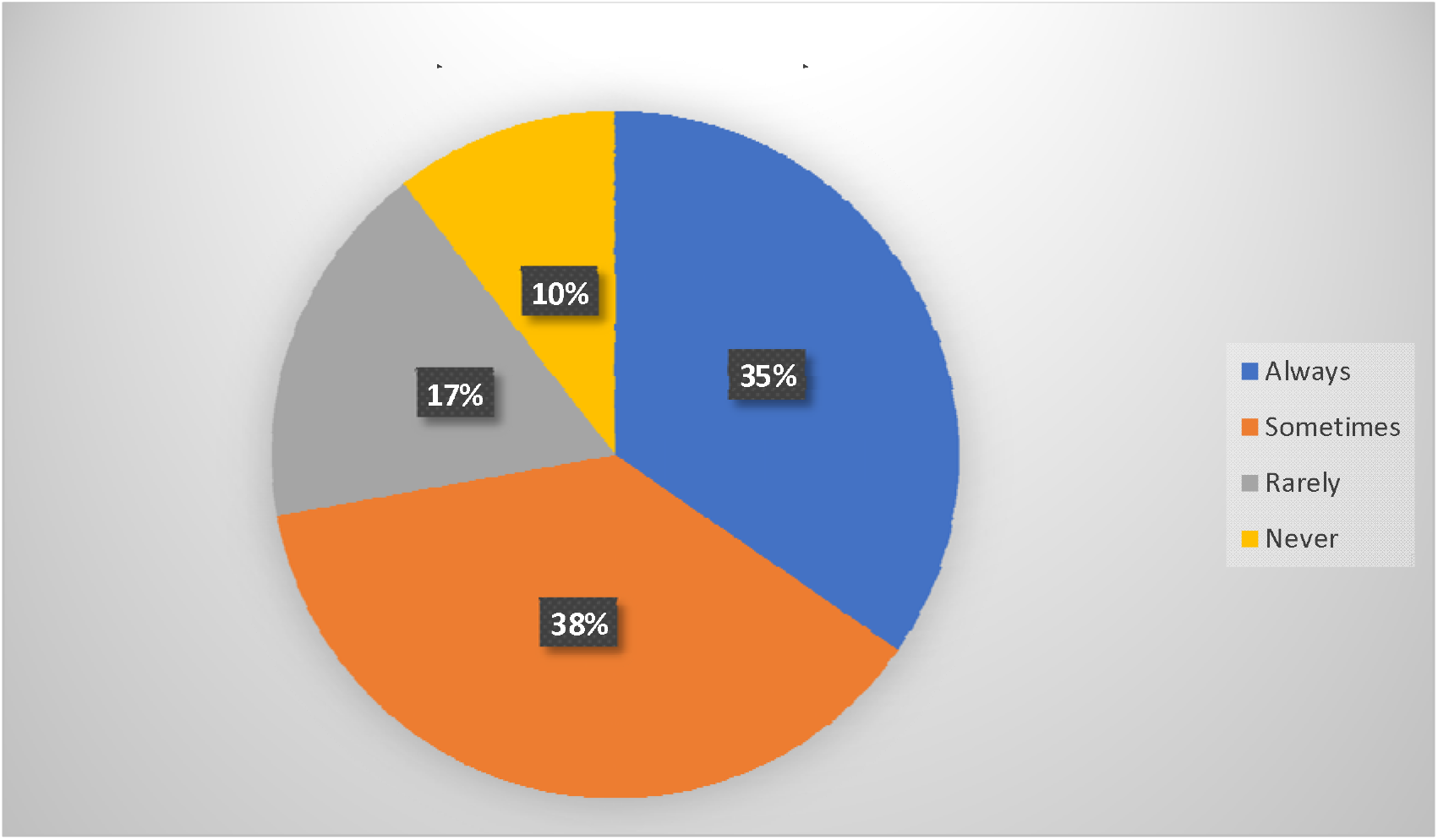

### Side Effects by Type (n=97)

Among the 97 participants who reported experiencing side effects from their anti-hypertensive medication, the most commonly reported side effects were dizziness (39.2%, n=38), frequent urination (24.7%, n=24), and cough (15.5%, n=15). Other side effects reported included headache (10.3%, n=10), swelling of the legs (5.2%, n=5), and nausea (5.2%, n=5). Some participants reported multiple side effects.

**Table:** Distribution of Side Effects by Type Among Participants Who Experienced Side Effects (n=97)

| Side Effect | Frequency (n) | Percentage (%)* |
| --- | --- | --- |
| Dizziness | 38 | 39.2 |
| Frequent urination | 24 | 24.7 |
| Cough | 15 | 15.5 |
| Headache | 10 | 10.3 |
| Swelling of legs | 5 | 5.2 |
| Nausea | 5 | 5.2 |

\*Percentages do not sum to 100% as participants could report multiple side effects.

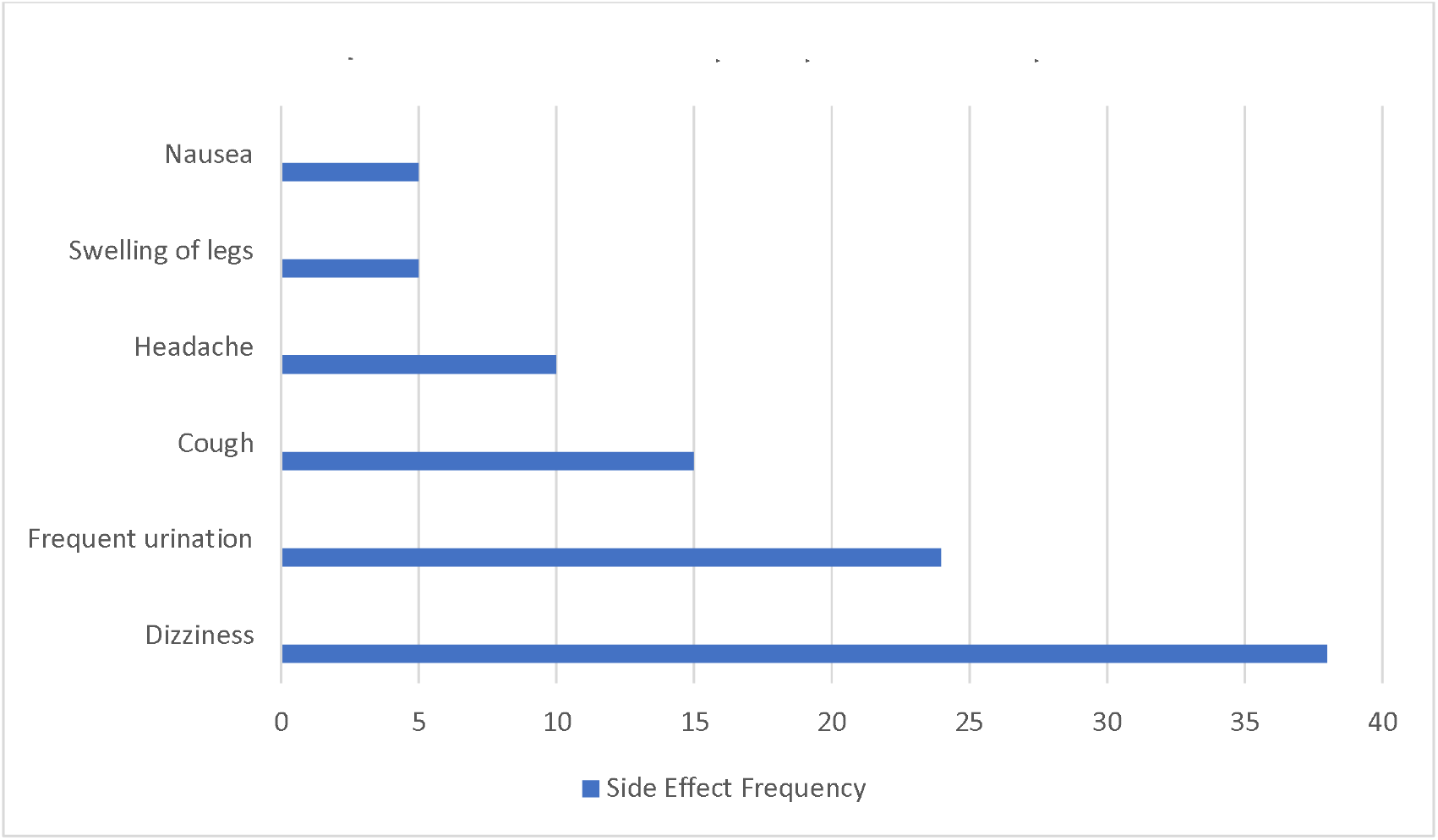

## DISCUSSION

### Statement of Principal Findings

This study investigated factors influencing adherence to anti-hypertensive medication among patients attending Railway Surgery Urban Health Centre in Ndola, Zambia. The findings reveal a substantial adherence challenge: only 41.4% of patients are adherent, with 58.6% classified as non-adherent. The most common reasons for non-adherence were running out of pills, being away from home, and forgetting, suggesting that most non-adherence was unintentional rather than intentional. Socio-demographic factors including age, education, and income, as well as clinical factors including comorbidities, side effects, and regimen complexity, were significantly associated with adherence. Health system barriers including transport problems, medication stock-outs, inadequate counselling, and out-of-pocket costs further compound the problem. These findings underscore the need for multi-level interventions addressing patient, provider, and health system factors.

### Level of Adherence to Anti-Hypertensive Medication

The finding of 41.4% adherence is concerning and may have significant implications for blood pressure control and cardiovascular outcomes in this patient population. The high rate of non-adherence (58.6%) suggests that many patients may not be receiving the full therapeutic benefit of their prescribed medications, potentially placing them at increased risk for complications such as stroke, heart failure, kidney disease, and premature mortality (Burnier & Egan, 2019; Abegaz et al., 2017).

The adherence rate found in this study is comparable to findings from other Zambian studies but shows some variation. Mapiki et al. (2023) reported a lower adherence rate of 35.7% among HIV patients on antihypertensive medication, which may reflect the compounded challenges faced by patients with multiple chronic conditions. Kampamba et al. (2021) found an adherence rate of 37.5% at Matero Level One Hospital in Lusaka, which is slightly lower than the current finding. In contrast, Mweene et al. (2010) reported a higher adherence rate of 70% among hypertensive patients at the University Teaching Hospital in Lusaka. These variations highlight the influence of different study populations, settings, methodologies, and adherence measurement tools, and underscore the importance of context-specific research.

Regionally, the adherence rate of 41.4% falls within the range reported in sub-Saharan African studies. Aminde et al. (2025) confirmed a high burden and increasing trend in non-adherence to blood pressure-lowering medications in their meta-analysis. Egekeze et al. (2025) found that implementation outcomes from adherence studies in Sub-Saharan Africa were suboptimal, with rates ranging from 30-50%. Globally, Abegaz et al. (2017) and Burnier & Egan (2019) reported that only about 50% of individuals follow their prescribed regimens, with adherence falling below 30% in some low-income settings (Kim et al., 2025; De Geest et al., 2021).

Analysis of individual Hill-Bone scale items revealed that the most common reasons for non-adherence were running out of pills (mean=2.6), being away from home (mean=2.5), and forgetting to take medication (mean=2.4). The least common reason was deciding not to take medication (mean=1.8), suggesting that most non-adherence was unintentional. This distinction is important because unintentional non-adherence may respond to interventions such as reminder systems, pill organisers, multi-month dispensing, and patient education about planning ahead for travel, while intentional non-adherence may require more intensive counselling and shared decision-making.

Mapiki et al. (2023) similarly found that practical barriers such as medication availability and access to healthcare facilities were significant contributors to non-adherence among HIV patients. This convergence of findings across different Zambian settings suggests that medication supply chain issues may be a widespread problem affecting adherence across the country.

### Socio-Demographic Factors Associated with Adherence Age

Age was significantly associated with adherence (p=0.006), with adherence rates increasing progressively from 21.4% among those aged 18-35 years to 50.0% among those aged above 65 years. In multivariate analysis, patients aged 51-65 years were nearly three times more likely to be adherent (AOR=2.89, 95% CI: 1.12-7.46), and those above 65 years were more than three times as likely to be adherent (AOR=3.24, 95% CI: 1.18-8.91) compared to those aged 18-35 years.

This finding is consistent with Mapiki et al. (2023) and Kim et al. (2025), who identified age as a consistent predictor of adherence. The association between younger age and poorer adherence may be related to competing demands from work and social activities, lower perceived vulnerability to complications, and more irregular daily routines. The better adherence observed among older patients may be related to established routines, greater experience with chronic disease management, and more realistic perceptions of health risks. Mweene et al. (2010) similarly found that age was significantly associated with adherence in their Lusaka study.

These findings suggest that interventions to improve adherence may need to be tailored to different age groups. For younger patients, reminder systems, flexible appointment scheduling, and educational messages emphasising long-term benefits may be beneficial. For older patients, regimen simplification and social support for medication management may be more appropriate.

### Education Level

Education was strongly associated with adherence (p=0.001), with rates increasing from 23.3% among those with no formal education to 57.4% among those with tertiary education. In multivariate analysis, patients with secondary education were nearly three times more likely to be adherent (AOR=2.84, 95% CI: 1.28-6.31), and those with tertiary education were more than three and a half times more likely to be adherent (AOR=3.56, 95% CI: 1.45-8.74) compared to those with no formal education.

This finding is consistent with Mapiki et al. (2023), Kampamba et al. (2021), and Kim et al. (2025), who identified education as a key predictor of adherence. Mweene et al. (2010) similarly found that patients with primary level education were more likely to be non-adherent. The relationship between education and adherence may be mediated by health literacy, understanding of hypertension as a chronic condition, and ability to navigate the healthcare system. Patients with lower education levels may benefit from simplified educational materials, more intensive counselling, and non-written communication methods such as pictures, demonstrations, and oral explanations in local languages.

### Monthly Income

Income showed a significant positive association with adherence (p=0.001), with rates increasing from 27.4% among those with no income to 75.0% among those earning K5,000 and above. In multivariate analysis, patients earning K5,000 and above were nearly six times more likely to be adherent (AOR=5.89, 95% CI: 1.92-18.07) compared to those with no income.

This finding aligns with Mapiki et al. (2023), who found that patients with limited financial resources were most affected by medication stock-outs. The mean monthly expenditure on medication and transport was K187.50 (SD=94.30), representing a significant burden for patients with no income (30.8%) or low income (26.2% earning less than K1,000). The association between income and adherence may be related to the ability to afford medications during stock-outs, access transportation, and manage competing basic needs. Interventions addressing financial barriers could include subsidising medications, ensuring consistent drug supply, providing transport vouchers, and exploring community-based medication delivery models.

### Non-Significant Socio-Demographic Factors

Sex (p=0.632), marital status (p=0.409), and employment status (p=0.177) did not show statistically significant associations with adherence. These findings are consistent with Kim et al. (2025), who noted that the relationship between socio-demographic factors and adherence is complex and context-dependent.

### Clinical Factors Associated with Adherence Presence of Comorbidities

Comorbidities were significantly associated with adherence (p=0.027), with patients having comorbidities showing lower adherence rates (33.3%) compared to those without (48.1%). In multivariate analysis, patients with comorbidities were 49% less likely to be adherent (AOR=0.51, 95% CI: 0.29-0.89). Among specific comorbidities, diabetes (p=0.042) and heart disease (p=0.025) were significantly associated with poorer adherence.

This finding is consistent with Mapiki et al. (2023) and Povia et al. (2023), who found that comorbidities were associated with poorer outcomes and adherence challenges. The presence of comorbidities may increase regimen complexity, treatment burden, and the likelihood of medication fatigue. Healthcare providers should screen for comorbidities, provide integrated care, minimise polypharmacy, and simplify regimens where possible.

### Experience of Side Effects

Side effects showed a strong association with adherence (p<0.001). Patients who experienced side effects had significantly lower adherence rates (26.8%) compared to those who did not (51.4%). In multivariate analysis, patients who experienced side effects were 64% less likely to be adherent (AOR=0.36, 95% CI: 0.20-0.65). This was the strongest negative predictor of adherence.

Mapiki et al. (2023) and Mweene et al. (2010) similarly found that side effects were associated with non-adherence. Healthcare providers should routinely inquire about side effects, take patient concerns seriously, and work collaboratively to find tolerable regimens. The finding that nearly half of participants (49.8%) received less than five minutes of counselling and 19.4% received no counselling at all suggests that current patient education practices may be inadequate for addressing side effect concerns.

The findings of this study are consistent with recent research in similar peri-urban Zambian settings. Mbewe et al. (2026) reported high discontinuation rates (43.2%) and missed appointments (51.5%) among injectable contraceptive users at Chipokota-Mayamba Clinic in Ndola, with side effects (54.4%) and fear of infertility (36.8%) as primary reasons for discontinuation—parallel to our findings where side effects were strongly associated with non-adherence (AOR=0.36, p=0.001). Similarly, Mapiki et al. (2026) found that only 17.8% of women at Masala Clinic in Ndola had adequate knowledge of intra-uterine devices, with 80.7% holding misconceptions about infertility.

Mapiki et al. (2026) further found a high prevalence of central obesity risk factors among adults in Kamakonde, Kitwe, with low awareness levels, mirroring our observation of limited health literacy as a barrier to adherence. These studies collectively highlight that method-specific fears, inadequate health knowledge, and health system challenges are cross-cutting barriers to adherence across both antihypertensive therapy and family planning in Zambia’s peri-urban settings.

### Medication Regimen Complexity

The number of pills taken per day was significantly associated with adherence (p=0.007). Adherence rates decreased from 55.2% for those taking one pill to 29.9% for those taking three or more pills daily. In multivariate analysis, patients taking three or more pills daily were 66% less likely to be adherent (AOR=0.34, 95% CI: 0.17-0.68).

This finding is consistent with the broader literature, including Kim et al. (2025), which identified regimen complexity as a consistent predictor of poor adherence. Where clinically appropriate, healthcare providers should prescribe fixed-dose combination pills, review regimens regularly to discontinue unnecessary medications, and provide adherence aids such as pill organisers and blister packs.

### Duration of Hypertension

Duration of hypertension diagnosis was not significantly associated with adherence (p=0.669). This finding is consistent with Kim et al. (2025), who found that disease duration was not consistently associated with adherence across studies, suggesting that other factors may be more important determinants.

### Health System Factors

Although not a specific objective, health system factors were assessed to provide context for adherence behaviour. The findings revealed several significant barriers that warrant discussion.

### Access to Care

The majority of participants (41.3%) lived 5-10 km from the clinic, and 21.1% lived more than 10 km away. Most (59.9%) relied on public transport, and over one-third (35.4%) reported missing an appointment due to lack of transport. These findings are consistent with Mapiki et al. (2023) and Chikumbu et al. (2024), who identified distance and transport as barriers to adherence. Addressing transport barriers may require decentralising services, implementing community-based medication distribution, providing transport vouchers, and multi-month dispensing.

### Medication Availability

Medication stock-outs were reported by 38.4% of participants in the past six months. This aligns with Mapiki et al. (2023) and Chikumbu et al. (2024), who identified stock-outs as a major health system barrier. Strengthening the drug supply chain, accurate forecasting, timely procurement, and contingency planning for shortages are essential. Improved communication with patients about stock-outs and expected resupply dates could help manage expectations.

### Patient-Provider Interaction

Nearly half of participants (49.8%) reported counselling time of less than five minutes, and 19.4% received no counselling at all. Only 34.6% felt providers always explained things clearly. These findings are concerning given the importance of effective communication in promoting adherence (Mapiki et al., 2023; Mweene et al., 2010). Interventions could include communication skills training for healthcare workers, use of local languages and visual aids, and allocation of adequate counselling time.

### Cost of Care

The mean monthly expenditure on medication and transport was K187.50 (SD=94.30), representing a significant burden for patients with no or low income. Muzambizi et al. (2021) noted that treatment adherence in Zambia is significantly affected by financial barriers. Ensuring consistent medication availability, exploring subsidies, and expanding health insurance coverage could reduce financial barriers.

### Integration of Findings with Conceptual Framework

The findings of this study align with the conceptual framework adapted from the World Health Organization’s Multidimensional Model of Adherence. The framework posits that adherence is influenced by the interplay of multiple factors, and the current study confirmed that socio-demographic factors (age, education, income) and clinical factors (comorbidities, side effects, regimen complexity) are independently associated with adherence. The finding that most non-adherence was unintentional suggests that interventions targeting practical barriers may be particularly effective. The high rate of non-adherence observed suggests that many patients may be at increased risk for poor blood pressure control and its complications, reinforcing the urgent need for interventions to improve adherence.

## WHAT IS ALREADY KNOWN ON THIS TOPIC

- Hypertension affects nearly 1.28 billion adults globally, with sub-Saharan Africa bearing a disproportionate burden (World Health Organization, 2017; Aminde et al., 2025).
- Non-adherence to anti-hypertensive medication ranges from 45-65% in sub-Saharan African settings (Aminde et al., 2025; Egekeze et al., 2025).
- In Zambia, adherence rates vary widely, with estimates ranging from 35.7% among HIV patients (Mapiki et al., 2023) to 70% in Lusaka (Mweene et al., 2010).
- Uncontrolled hypertension contributes substantially to hospital morbidity and mortality in Zambia (Povia et al., 2023; Muzambizi et al., 2021).

## WHAT THIS STUDY ADDS

- Adherence to anti-hypertensive medication at Railway Surgery Urban Health Centre is suboptimal, with only 41.4% of patients classified as adherent.
- Most non-adherence is unintentional, driven by running out of pills, being away from home, and forgetting, rather than intentional decisions to stop treatment.
- Age, education, income, comorbidities, side effects, and regimen complexity are independently associated with adherence in this population.
- Health system barriers including transport problems, medication stock-outs, and inadequate patient-provider communication compound adherence challenges.

## CONCLUSION

This study assessed the factors associated with adherence to anti-hypertensive medication among patients attending Railway Surgery Urban Health Centre in Ndola, Zambia. The findings reveal that adherence is suboptimal, with only 41.4% of patients classified as adherent. The majority of non-adherence is unintentional, suggesting that practical interventions could have substantial impact. Socio-demographic factors including age, education, and income, as well as clinical factors including comorbidities, side effects, and regimen complexity, are significantly associated with adherence. Health system barriers including transport problems, medication stock-outs, inadequate counselling, and out-of-pocket costs further compound the problem. Addressing these barriers requires a multi-level response including regimen simplification, proactive side effect management, tailored patient education, medication supply chain strengthening, transport support, and enhanced patient-provider communication. Without such targeted efforts, the escalating burden of uncontrolled hypertension at Railway Surgery Urban Health Centre is likely to persist, with consequences for individual patients, the healthcare system, and society.

## STRENGTHS AND LIMITATIONS

### Strengths

This study has several strengths. First, the use of a validated and modified Hill-Bone Compliance Scale, adapted from Mweene et al. (2010), provided a standardised and reliable measure of adherence that has been previously used in the Zambian context, enhancing comparability with other local studies. Second, the sample size of 237 participants, calculated using Cochran’s formula, provided adequate statistical power to detect associations between multiple independent variables and adherence status, with an events-per-variable ratio of 16.3 exceeding the recommended minimum of 10. Third, the use of simple random sampling from the clinic’s patient register minimised selection bias and enhanced the representativeness of the sample. Fourth, the study employed both bivariate and multivariable analytical methods, allowing for the identification of factors independently associated with adherence while controlling for potential confounders. Fifth, the inclusion of health system factors provided a comprehensive understanding of adherence barriers beyond individual-level characteristics, offering insights for multi-level interventions. Sixth, the high response rate of 93% minimised non-response bias and enhanced the generalisability of findings to the clinic population. Seventh, the study focused on a public primary healthcare facility, providing evidence that is relevant to similar settings across Zambia and the region. Eighth, the use of interviewer-administered questionnaires with trained research assistants and translation into the local language (Bemba) ensured comprehension and accurate data collection across diverse literacy levels.

## Limitations

Several limitations should be considered when interpreting these findings. First, the cross-sectional design precludes causal inference; the associations identified should be interpreted as correlates rather than determinants of adherence. Second, reliance on self-reported adherence may be subject to social desirability bias, as patients may overreport adherence due to perceived expectations. However, the modified Hill-Bone scale has been validated and used in similar settings, and the consistency of findings with objective measures from other studies supports their validity. Third, the study was confined to one urban health centre in Ndola, which limits the generalisability of findings to other settings in Zambia, particularly rural areas where access to care and health system resources may differ. Fourth, the study did not include objective measures of adherence such as pill counts, pharmacy refill records, or biochemical markers, which could have provided more accurate adherence estimates. Fifth, the study did not collect data on household income in a standardised manner, and the monthly income categories used may not fully capture economic barriers to adherence. Sixth, the study did not include qualitative components to explore patients’ lived experiences and perceptions in depth, which could have provided richer insights into the barriers and enablers identified. Seventh, the study did not assess the effect of adherence on clinical outcomes such as blood pressure control, which would have strengthened the clinical relevance of the findings. Eighth, the sample size, while adequate for overall analysis, may be underpowered for some subgroup analyses, limiting the ability to detect differences between specific groups. Ninth, the study did not include healthcare provider perspectives, missing an opportunity to understand systemic barriers from the provider’s viewpoint.

## RECOMMENDATIONS

Based on the findings of this study, the following recommendations are made:

### 1. Strengthen the Drug Supply Chain to Prevent Medication Stock-Outs

Given that 38.4% of participants reported medication stock-outs in the past six months and running out of pills was the most common reason for non-adherence (mean=2.6), the clinic management should strengthen the drug supply chain through accurate forecasting, timely procurement, adequate storage, and contingency planning for shortages. Multi-month dispensing for stable patients should be implemented to reduce visit frequency and buffer against stock interruptions.

### 2. Proactively Identify and Manage Medication Side Effects

Given that patients who experienced side effects were 64% less likely to be adherent (AOR=0.36, 95% CI: 0.20-0.65), healthcare providers should routinely inquire about side effects at each visit, take patient concerns seriously, and work collaboratively to find tolerable alternatives. This may involve adjusting doses, switching to different medication classes, or providing strategies to manage side effects without discontinuing medication. Patients should be counselled about potential side effects when medications are initiated.

### 3. Simplify Medication Regimens Where Clinically Appropriate

Given that patients taking three or more pills daily were 66% less likely to be adherent (AOR=0.34, 95% CI: 0.17-0.68), healthcare providers should prescribe fixed-dose combination pills where clinically appropriate, conduct regular medication reviews to discontinue unnecessary medications, and simplify dosing schedules. Adherence aids such as pill organisers, blister packs, and reminder systems should be provided for patients on complex regimens.

### 4. Provide Tailored Patient Education that Considers Literacy Levels

Given that patients with no formal education were 3.5 times less likely to be adherent compared to those with tertiary education (AOR=3.56, 95% CI: 1.45-8.74), health education should be tailored to patients’ literacy levels using simple, non-written materials, pictures, demonstrations, and oral explanations in local languages. The teach-back method should be used to verify understanding. Emphasis should be placed on the chronic nature of hypertension, the importance of adherence even when feeling well, and practical strategies for remembering medication.

### 5. Address Financial Barriers to Care

Given that patients earning K5,000 and above were nearly six times more likely to be adherent compared to those with no income (AOR=5.89, 95% CI: 1.92-18.07), and the mean monthly expenditure on medication and transport was K187.50, interventions to reduce financial barriers should be explored. These could include subsidising medications for low-income patients, providing transport vouchers or reimbursement, eliminating user fees for chronic disease care, and exploring community-based medication delivery models.

### 6. Enhance Patient-Provider Communication and Counselling

Given that nearly half of participants (49.8%) received less than five minutes of counselling, 19.4% received no counselling at all, and only 34.6% felt providers always explained things clearly, the clinic should allocate adequate time for patient counselling and provide communication skills training for healthcare workers. Training should emphasise explaining complex medical information in simple terms, using local languages, checking patient understanding, and addressing patients’ anxieties and concerns about medication.

### 7. Implement Targeted Interventions for Younger Patients and Those with Comorbidities

Given that younger patients (18-35 years) had the lowest adherence rates (21.4%) and patients with comorbidities were 49% less likely to be adherent (AOR=0.51, 95% CI: 0.29-0.89), specific interventions should target these high-risk groups. For younger patients, strategies might include mobile phone reminders, flexible appointment scheduling, peer support groups, and educational messages emphasising long-term benefits. For patients with comorbidities, integrated care models, comprehensive medication reviews, and coordinated care across specialties should be implemented to reduce treatment burden.

## Data Availability

All data produced in the present study are available upon reasonable request to the authors

